# Implementation of a Comprehensive Telehealth Assessment Battery for Complex Neurogenetic Disease: An Observational Study of Rapid-Onset Dystonia-Parkinsonism

**DOI:** 10.64898/2026.02.18.26345928

**Authors:** Ihtsham U Haq, Daniel Sirica, Vicki L. Wheelock, Ralph H Benedict, Marina L Sarno, Kris Tjaden, Laurie J Ozelius, Rebecca Firth, Eleonora Napoli, Kathleen J Sweadner, Allison Brashear

## Abstract

ATP1A3-related syndromes represent a continuously expanding clinical spectrum and present with an extraordinarily wide range of symptoms. New phenotypes continue to emerge, posing ongoing challenges for both diagnosis and development of treatments. In this context, telemedicine offers a unique opportunity to greatly expand outreach to patients. Remote, high-resolution assessments help refine phenotypic characterization and the identification of novel and intermediate phenotypes.

In this study we aimed to determine completion rates and practicality of conducting motor, speech, and neuropsychological assessments entirely via virtual visits. Although the broader recruitment included several ATP1A3-related disorders, this virtual battery was specifically developed for subjects with RDP. Participants with other ATP1A3 phenotypes enrolled in the study contributed to evaluating the overall feasibility of the workflow but were not the target population for the full battery.

We recruited individuals with suspected or confirmed diagnosis of ATP1A3-related disorders, along with familial controls, from three participant clinical sites. Participants completed all study procedures through scheduled telemedicine visits using their personal devices (tablets, laptops, smartphones). A total of 59 participants were enrolled, including 46 individuals with suspected or confirmed ATP1A3 variants and 13 family member controls. Among affected patients, 18 had RDP, 12 AHC (Alternating Hemiplegia of Childhood), 4 CAPOS (Cerebellar ataxia, Areflexia, Pes cavus, Optic atrophy, Sensorineural hearing loss), 10 were categorized as “uncertain” and 2 with “mixed” phenotype (RDP/CAPOS and RDP/AHC).

The virtual assessment battery included: (i) patient history questionnaire (PHQ), (ii) structured neurological examination adapted for virtual visits, (iii) speech recording, and (iv) extensive neuropsychological assessment. Feasibility was evaluated based on completion rates for each assessment component.

Remote neurological, speech and neurocognitive/psychiatric assessments were completed by most participants with ATP1A3 phenotypes, with completion rates of 78% for motor examination and 87% for speech evaluation. The observed pattern of motor and speech impairments were consistent with prior in-person evaluations, supporting the validity and feasibility for both motor and nonmotor features of remote assessment in complex genetic neurological disease.

## INTRODUCTION

Rapid-Onset Dystonia-Parkinsonism (RDP) is a rare ATP1A3-related disorder characterized by abrupt onset of dystonia, parkinsonism, bulbar dysfunction, and gait impairment, often precipitated by stressors ^1^. Since our original 1993 description of RDP in a single family ^2^, it has become increasingly evident that the clinical presentation of this disorder is more heterogeneous and anatomically dispersed than previously believed ^3–6^. Cognizant of this, in recent years, our interdisciplinary team (clinical, genetics, neuropathology, imaging, cell biology and biochemistry) adopted a strategy to better understand ATP1A3-related disorders, with a special focus on RDP.

The heterogeneity of RDP symptoms makes thorough evaluation essential for accurate diagnosis and timely symptom management. Especially in the case of rare neurological disorders, swift access to specialized experts remains a major barrier for many patients. Patients with RDP are scattered across the United States and often lack access to clinical centers or neurologists with expertise in movement disorders. Furthermore, many face challenges travelling due to physical disability and/or the financial burden associated with travel to specialized medical facilities.

“Teleneurology” ^7^ represents a rapidly evolving area of telemedicine focused on delivering neurological care remotely and digitally, which in recent years has been successfully applied to a wide range of neurological conditions, including headache, dementia, epilepsy, concussion/TBI, stroke, movement disorders, and multiple sclerosis ^8^. Recent advances in telemedicine tools, standardized virtual examination protocols, and remote monitoring techniques have favored the expansion of teleneurology to longitudinal disease assessment and management as well as phenotyping ^9,10^.

Here, we present a comprehensive multidomain assessment battery (neurological, cognitive, psychiatric and speech measures) that can be successfully administered via telemedicine, incorporating new testing criteria designed to remove barriers to diagnosis and to facilitate development of targeted treatments. The implementation of this fully remote-assessment protocol enabled characterization of patients within their own familiar environment, expanding both outreach and breadth of ATP1A3-related research. This represents a significant advance in our ability to quantify the symptoms and phenotypic spectrum characterizing RDP. Key features include: (i) higher-resolution quantitation of motor dysfunction; (ii) blinded, history-independent neurological examination; (iii) a tailored neurocognitive and psychiatric approach using scales designed to detect subtle, age-dependent manifestations of psychiatric illness and cognitive impairment; and (iv) systematic assessment of speech production abnormalities, a practical and portable indicator that could be utilized in future clinical trials.

We believe that leveraging telemedicine and emerging digital technology, whose reach and sophistication have grown exponentially in recent years, will help refine our understanding of ATP1A3 mutation phenotype diversity. Remote assessment not only facilitates broader patient participation but also enables the collection of real-world data on motor, behavioral and cognitive function. Integration of these data with genetic and imaging findings will advance our understanding of ATP1A3 phenotypic diversity, providing a more complete picture of disease progression across the lifespan.

## METHODS

### Study design and subjects

To ensure broad geographic representation and to evaluate feasibility of remote assessment across diverse settings, this prospective longitudinal observational study was conducted across three participating sites representing major US regions: the University at Buffalo (UB, Northeast), the University of Miami (UM, East Coast), and the University of California, Davis (UCD, West Coast). This multisite design enabled recruitment from distinct geographic areas and testing of the telemedicine framework across varied technological environments and patient populations.

We enrolled a total of 68 participants (41 females and 27 males; **Table 1**), of whom 59 underwent a completely virtual assessment (9 participants underwent in-person evaluation and were not included in this study; **Figure 1**). Among these, 46 were ATP1A3-mutation positive, or had a clinically suspected ATP1A3 phenotype, including 18 individuals with RDP, 12 with AHC, 4 with CAPOS syndrome, and 2 with “mixed” phenotype (one RDP/CAPOS, and one RDP/AHC). Ten participants (hereafter referred to as having “uncertain phenotype”) had an uncertain diagnosis or variant with unknown significance. The remaining 13 participants were controls recruited from families of mutation-positive subjects.

**Figure 1.**
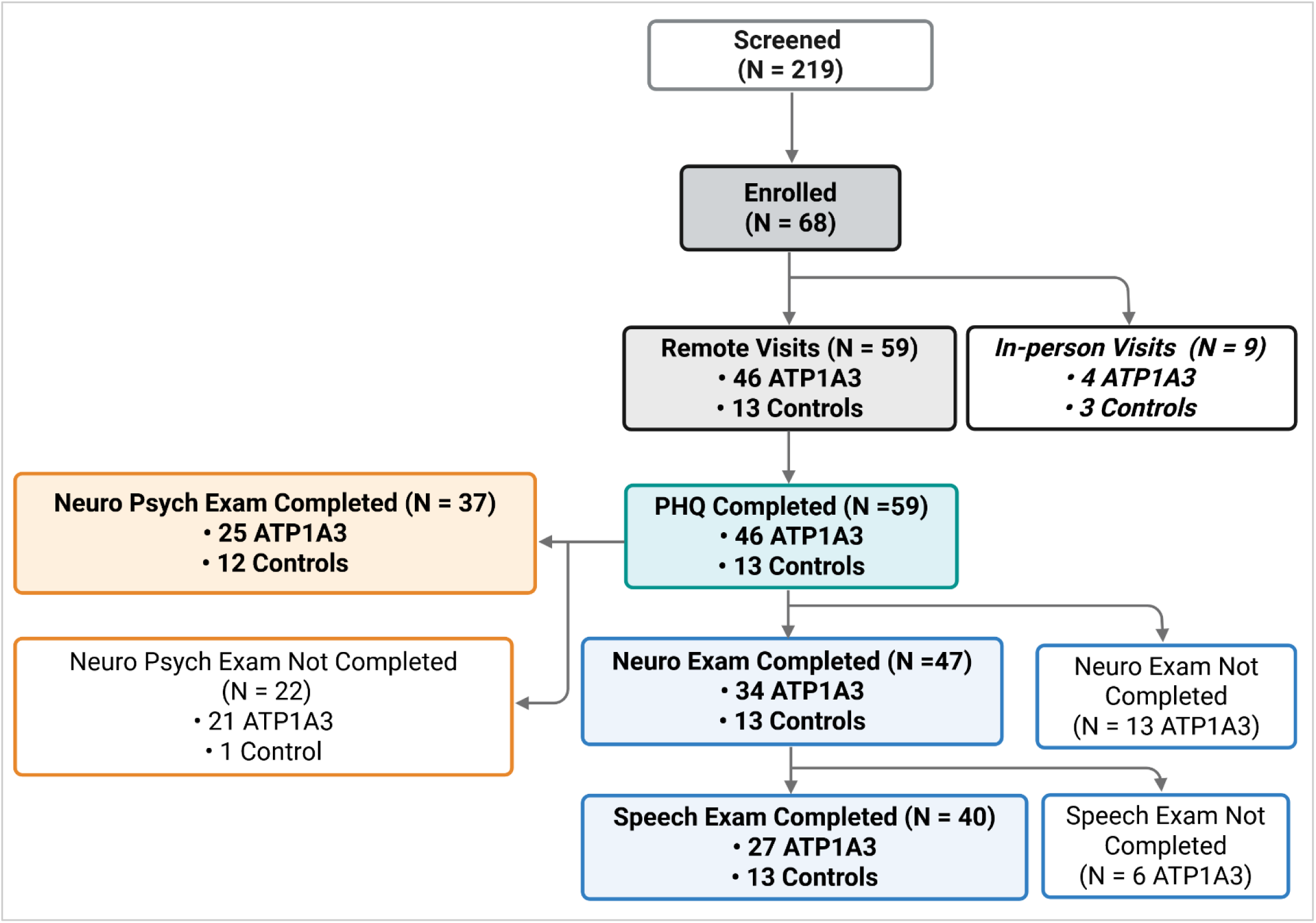
Participant enrollment and assessment workflow. Flowchart illustrating the progression of participants through the different stages of the study, including screening, enrollment, format, and completion of assessments. Of 219 individuals screened, 68 were enrolled either for remote (n = 59) or in-person visits (n = 9). Completion rates for PHQ, neurological examinations, speech evaluations, and neurocognitive/neuropsychological assessments are also shown with correspondent number of participants with ATP1A3 variants and controls at each stage. Flowchart was created with BioRender.com.

**Table 1.** Participant Demographics.

|  | **N (%)** | **Age y** | **Age of Onset** |
| --- | --- | --- | --- |
| **Familial controls** (8 females, 5 males) | 13 | 27.2 ± 15.6<br>[7, 63] | - |
| **ATP1A3-related diagnoses** (26 females, 20 males) | 46 | 23.1 ± 17.3<br>[1, 59] | 9.7 ± 13.9<br>[0.0, 56.0] |
| • **RDP** | 18 (39.1) | 32.5 ± 18.0<br>[7, 59] | 20.7 ± 15.5<br>[4.3, 56.0] |
| • **AHC** | 12 (26.0) | 21.0 ± 14.3<br>[1, 49] | 1.8 ± 4.5<br>[0.0, 16.0] |
| • **CAPOS** | 4 (8.7) | 22.3 ± 18.2<br>[10, 49] | 3.1 ± 3.3<br>[1.3, 8.0] |
| • **Mixed phenotype\*** | 2 (4.3) | - | - |
| • **Undefined phenotype** | 10 (21.7) | 11.4 ± 12.6<br>[1-57] | 1.1 ± 1.9<br>[0.0, 6.0] |
| **Subjects with ATP1A2 variants** | 2 | 7.0 ± 5.7 | - |
| **Participants With a Follow-Up Visit To Date** | 19 (27.9) | - | - |

\*Two subjects with RDP were diagnosed with an additional phenotype of respectively CAPOS and AHC. Data are shown as mean ± SD, and interval [min, max]

Age at symptom onset (**Table 1**) varied substantially across self-reported ATP1A3 diagnostic subgroups. In the overall ATP1A3 cohort mean age of onset was 9.7 (± 13.9; SD) years, with a median of 1.7 and a wide range extending into adulthood, reflecting wide phenotypic heterogeneity. Not surprisingly, individuals with self-reported AHC and CAPOS exhibited the earliest onset, with median ages of 0.4 and 1.6 years, respectively, consistent with predominantly early-childhood presentations. Conversely, participants with self-reported RDP demonstrated substantially later onset, with mean age of 20.7 (± 15.5) years and median of 15.5 years, reflecting adolescence and adult disease onset. Participants with uncertain phenotypes showed earlier onset overall, with a mean age of 1.1 (± 1.9) years. There can be a distinction between self-reported diagnosis and diagnosis by criteria; our group has adhered to a definitional division between AHC and RDP as motor onset before (AHC) or after (RDP) 18mo, in the presence of an ATP1A3 variants ^11^.

Participants underwent initial virtual assessment as part of our ongoing longitudinal protocol, which aimed to evaluate the feasibility of conducting neurological, speech/voice and neuropsychological assessment entirely via telemedicine. Of those enrolled in the telemedicine cohort, 47 individuals (13 controls and 34 ATP1A3 cases) completed at least one neurological visit using secure, HIPAA-compliant telemedicine platforms (**Figure 1**).

### Telemedicine setup, infrastructure and testing materials

All study procedures were conducted using a fully remote telemedicine framework designed to support secure, standardized assessments across geographically dispersed sites. This decentralized setup enabled consistent administration of the protocol across three geographically distributed sites (East Coast, Northeast, and West Coast), and allowed individuals with limited mobility or high travel burden to participate fully from their homes. Prior to their scheduled telehealth session, participants (or their caregivers, when applicable) were mailed a standardized testing kit containing the materials required for remote assessment. Kits included: a measuring tape for spatial measurements during the neurological exam; pre-measured distance ropes for the Timed Up and Go, and 4- and 10-meter walk tasks; a plastic straw for cranial nerve V assessment; a Zoom H1 digital audio recorder for high-quality speech and voice sampling; printed reading passages for standardized speech elicitation; and a saliva collection tube with accompanying packaging material for sample return. Instructions for use were included with each kit.

Virtual visits were conducted through HIPAA-compliant videoconferencing platforms approved by each participating institution (Webex or Zoom for telehealth). Participants connected using personal devices (smartphone, tablet, or laptop computer depending on device availability), with no additional hardware required. All device types provided adequate video and audio quality for examination, with laptop offering greater flexibility in camera positioning and tablets providing more user-friendliness for caregiver-assisted participants. Video and audio quality were sufficient for blinded ratings.

Examiners followed a standardized protocol for camera positioning, lighting, audio optimization, and participant/caregiver guidance. For younger participants or those with significant motor impairment, caregivers were instructed to assist with camera stabilization, task execution, and handling of materials.

Participants received verbal instructions before the session on how to prepare the testing space, set up the camera, enable adequate WIFI/internet connection, and ensure a quiet distraction-free environment. At the start of each visit, examiners confirmed connectivity, tested audio and video settings, and guided participants through troubleshooting when needed.

All digital data, including questionnaires, neurological ratings, speech audio recordings, and neuropsychological raw data were uploaded to secure institutional servers following each visit, with no data stored on participant devices.

### Telemedicine-based clinical phenotyping battery

All participants (n = 59) completed a multimodal assessment battery that included: (i) a telemedicine-adapted Patient History Questionnaire (PHQ; **Table 2**); (ii) a family history form (**Table 3**); (iii) a structured neurological examination (**Table 4**) modeled on protocols developed and approved by the Dystonia Coalition and adapted for virtual visits; (iv) a speech and voice assessment protocol (**Table 4**); and (v) a comprehensive neuropsychological evaluation appropriate for remote testing (**Table 5**).

**Table 2.** Domains and Outcomes in the Patient History Questionnaire (PHQ)

| **Domain** | **Content / Outcomes Collected** |
| --- | --- |
| **I. Demographics & Background** | Family ID, parental names/ages, education level, independent living status, ancestry, gender identity, handedness, height. |
| **II. Neurological & Motor History** | Eye movement abnormalities; facial, jaw, tongue dystonia; swallowing difficulties; drooling; speech effort; head/neck deviation; tremors; limb dystonia; clumsiness; cramps; numbness/tingling; scoliosis; functional motor limitations. |
| **III. Seizure History** | Lifetime seizures, suspicious spells, seizure types (absence, tonic-clonic, focal, myoclonic, tonic, atonic), age at onset, and frequency. |
| **IV. Headache/Migraine History** | Occurrence, recent episodes, associated nausea/photophobia, functional impact, prior MRI/EEG. |
| **V. Hearing Problems** | Difficulties hearing in various contexts, social withdrawal, tinnitus, dizziness, pain. |
| **VI. Mood & Psychiatric History** | Depression, anhedonia, bipolar symptoms, anxiety disorders; diagnoses, treatments, and contextual factors contributing to symptoms. |
| **VII. Developmental History** | Pregnancy/delivery complications, maternal exposures, developmental milestones (walking/talking), school difficulties, learning disabilities, special services (speech/OT/PT), behavioral issues. |
| **VIII. ATP1A3 Diagnosis** | Confirmed or suspected ATP1A3 disorders (RDP, AHC, CAPOS), age at symptom onset, age at diagnosis, diagnosing clinician and location. |
| **IX. Current Symptoms (Last 30 Days)** | Body-region-specific presence of symptoms (face, mouth, arms, legs, neck, trunk), and changes over past year and since onset. |
| **X. Symptom Course Over Time** | Patient-selected course chart describing trajectory; examiner-classified course (single episode, relapsing/remitting, relapsing/progressive, primary progressive). |
| **XI. Onset Characteristics** | Speed of onset (minutes → months), initial body region, early cramps, tremor or pain at onset, stabilization time, periods of remission. |
| **XII. Trigger-Related Factors** | Physical exertion: 1 month → 24h prior.<br>Life stressors: 1 month → 24h prior.<br>Heat exposure or fever: 1 month → at onset.<br>Infections: 1 month → at onset. |
| **XIII. Alcohol Use History** | Drinking patterns: 24h → 1 year prior; number of drinks on typical and peak days; intoxication frequency. |
| **XIV. Other Potential Triggers** | Open-ended description of any additional triggering factors. |
| **XV. Medication History & Treatment Response** | Detailed trial history across: cardiac meds, Parkinson's meds, anticholinergics, other medications, supplements. Each includes: use (yes/no), benefit (yes/no), degree of benefit (slight/moderate/complete). |
| **XVI. Additional Notes & Data Collection** | Free-text comments; informant involvement; mode of data collection (in-person, virtual, video review, self-administered). |

**Table 3.** Domains and Outcomes Collected in the Family History Form.

| **Domain** | **Outcomes Collected** |
| --- | --- |
| **I. Informant Information** | Proband ID, Informant ID, Acrostic, Date of collection, Informant's relationship to proband, rater scores for reliability (1–5), willingness (1–5), truthfulness (1–5). |
| **II. Basic Demographics (per family member)** | Individual ID, first name, year of birth, biological father ID, biological mother ID, gender (M/F/DK), deceased (Y/N/DK), age and cause of death if deceased. |
| **III. Family Structure Variables** | Multiple birth status (identical/fraternal/DK/N/A), adopted status (Y/N/DK). |
| **IV. Core Neurological Features** | Dystonia (Y/N/DK + age of onset), Parkinsonism (Y/N/DK + age of onset), RDP (Y/N/DK + age of onset). |
| **V. ATP1A3-Related Phenotypes** | AHC (Y/N/DK + age of onset), CAPOS (Y/N/DK + age of onset). |
| **VI. Additional Neurological Conditions** | Migraines, seizures, deafness (each: Y/N/DK). |
| **VII. Psychiatric Symptoms** | Depression, anxiety, psychosis, suicide attempts (each: Y/N/DK). |
| **VIII. Neurodevelopmental & Behavioral Features** | Learning problems, behavior problems, anger management problems, ADHD (each: Y/N/DK). |
| **IX. Genetic Testing** | Whether genotyped (Y/N/DK), test result (positive/negative). |

**Table 4.** Neurological Exam Session Description.

| Step | Section / Task | Subtasks |
| --- | --- | --- |
| 0 | Pre-Visit Tasks | Intake, Contact Info, Demographics, Family History; PHQ; CPIB; VHI-10 |
| 1 | Voice & Speech Assessment | Recorder setup → sustained vowels → AMR tasks, syllable repetition (puh/tuh/kuh) → SMR (puhtuhkuh) → counting 1–10 → reading (Caterpillar, Rainbow, SIT) → Apraxia/Dysarthria (<12 yrs) → EAT-10 + swallowing questions |
| 2 | Cranial Nerves III/IV/VI | Horizontal/vertical gaze → fast saccades → gaze-evoked nystagmus → ocular pursuit → saccade dysmetria |
| 3 | Cranial Nerve V | Facial light touch → bite strength R/L |
| 4 | Cranial Nerve VII | Forehead wrinkle → eye closure → cheek puff |
| 5 | Cranial Nerve VIII | Finger rub hearing tests |
| 6 | Cranial Nerve XI | Shoulder shrug |
| 7 | Cranial Nerve XII | Tongue protrusion → tongue lateralization → mouth opening dystonia (BFMDRS 2) |
| 8 | Upper Face & Eyes | UPDRS 1.2 facial expression → BFMDRS (1) eye dystonia |
| 9 | Pour–Drink Test | Water pouring R/L → Drink test R/L |
| 10 | Neck | Neutral rest (eyes open/closed) → rotation R/L → lateral flexion → flex/extend → lateral view |
| 11 | Arms (postures and function) | Arm postures (supinated/pronated/flexed) → finger tapping R/L → wrist flex/ext → cup-to-lips R/L (BFMDRS 5–6) |
| 12 | Fine Motor: UPDRS Hand Series | UPDRS 1.4 finger tapping → 1.5 hand opening/closing → 1.6 pronation–supination |
| 13 | Tremor Assessments | UPDRS 1.15 postural tremor → 1.16 kinetic tremor → ICARS 10–12 (finger–nose, intention tremor, finger–finger) |
| 14 | Writing & Spirals | Write sentence ×3 → freehand spirals R/L → Archimedes spirals R/L |
| 15 | Trunk & Lower Body | ICARS 7 sitting → BFMDRS trunk exam standing & walking → heel–toe tapping R/L → ICARS knee–tibia tests (8–9) |
| 16 | Lower Limb UPDRS Series | Toe tapping (UPDRS 1.7) R/L → Leg agility (UPDRS 1.8) R/L |
| 17 | Gait Assessment Part I | UPDRS 1.9 arising → UPDRS 1.10 gait → UPDRS 1.11 freezing |
| 18 | Gait Assessment Part II | UPDRS 1.13 posture → ICARS 1–6 (walking capacity, gait speed, stance, sway) → UPDRS 1.14 global bradykinesia |
| 19 | Rest Tremor Assessments | UPDRS 1.17 amplitude (all 4 limbs + jaw) → UPDRS 1.18 constancy |
| 20 | Medication Status | UPDRS 1.a–c Parkinson’s meds → levodopa timing |
| 21 | Functional Mobility Tests | 4-meter walk (practice + 2 trials) → TUG (practice + 2 trials) |
| 22 | End of Recording | Participant holds ID/date sign for 5 seconds |

**Table 5.** Cognitive and Neuropsychological Measure.

| **Measure** | **Domain Assessed** |
| --- | --- |
| **Reynolds Intellectual Assessment Scales (RIAS) – Odd-Item** | Nonverbal Reasoning / Fluid Intelligence |
| **NEPSY-II Comprehension of Instructions** | Auditory Comprehension / Receptive Language |
| **Test of Visual Perceptual Skills (TVPS) – Sequential Memory** | Visual Sequential Memory / Visual Working Memory |
| **WRAML-2 Verbal Learning** | Verbal Learning / Episodic Memory |
| **Benson Figure Copy** | Visuospatial Construction / Visual Perception |
| **Oral Trail Making Test (Oral TMT)** | Executive Function / Set-Shifting / Processing Speed |
| **Symbol Digit Modalities Test (SDMT)** | Processing Speed / Attention |
| **WRAML-2 Verbal Learning Delayed Recall** | Verbal Memory – Delayed Recall |
| **WRAML-2 Verbal Learning Recognition** | Verbal Memory – Recognition |
| **Benson Figure Delayed Recall** | Visual Memory – Delayed Recall |
| **COWAT Verbal Fluency Test** | Executive Function / Language Fluency |
| **Mood Assessment Scale (MAS) – GDS** | Mood / Geriatric Depression |
| **Hamilton Depression Rating Scale (HDRS)** | Mood – Depression Severity |
| **Children’s Depression Inventory-2 (CDI-2)** | Mood – Childhood Depression |
| **Hamilton Anxiety Rating Scale (HAM-A)** | Anxiety |
| **Yale-Brown Obsessive Compulsive Scale (Y-BOCS)** | Obsessive-Compulsive Symptoms |
| **Lawton–Brody Instrumental Activities of Daily Living (IADL)** | Functional Independence / Daily Living Skills |
| **Quick SCID-5** | Psychiatric Diagnostic Interview / Clinical Diagnosis |
| **WRAML-2 Story Memory (≤8 yrs, Stories A & B)** | Verbal Episodic Memory – Immediate Recall |
| **WRAML-2 Story Memory (≥9 yrs, Stories B & C)** | Verbal Episodic Memory – Immediate Recall |
| **WAIS-IV Digit Span** | Attention / Working Memory |
| **WISC-IV Digit Span (6–16 yrs)** | Attention / Working Memory |
| **WAIS-IV Matrix Reasoning** | Fluid Reasoning / Visual Abstract Reasoning |
| **WISC-V Matrix Reasoning (6–16 yrs)** | Fluid Reasoning / Visual Abstract Reasoning |
| **WRAML-2 Story Memory Delayed Recall (≤8 yrs)** | Verbal Memory – Delayed Recall |
| **WRAML-2 Story Memory Recognition (≤8 yrs)** | Verbal Memory – Recognition |
| **WRAML-2 Story Memory Delayed Recall (≥9 yrs)** | Verbal Memory – Delayed Recall |
| **WRAML-2 Story Memory Recognition (≥9 yrs)** | Verbal Memory – Recognition |
| **WAIS-IV Vocabulary** | Language / Semantic Knowledge |
| **WISC-V Vocabulary (6–16 yrs)** | Language / Semantic Knowledge |
| **WAIS-IV Similarities** | Verbal Reasoning / Abstract Thinking |
| **WISC-V Similarities (6–16 yrs)** | Verbal Reasoning / Abstract Thinking |
| **Boston Naming Test (BNT)** | Confrontation Naming / Language |
| **WCST-64 (online)** | Executive Function / Cognitive Flexibility / Problem-Solving |
| **BRIEF-2 (Parent)** | Everyday Executive Function – Children |
| **BASC-3 (Parent)** | Behavioral & Emotional Functioning – Children/Adolescents |
| **BRIEF-A (Adult)** | Everyday Executive Function – Adults |

The order of tests presented here reflects the order of administration.

All examiners were trained clinicians familiar with both in-person and remote neurological and neuropsychological assessments and certified on the component rating scales where certification was possible. Each session was recorded (via Zoom or Webex) and reviewed by the four study neurologists during regular videoconference consensus meetings to ensure consistent interpretation across different sites. During these sessions, examiners jointly reviewed selected recordings, discussed scoring decisions and resolved discrepancies.

#### History questionnaires (PHQ and Family History Form)

The PHQ and family history (**Tables 2** and **3**) were administered before the neurological examination and designed to assess historical and phenomenological associations with patient symptoms.

The PHQ is a comprehensive (**Table 2**), 279-item instrument designed to capture detailed medical, developmental, psychiatric, genetic, and symptom-specific information in individuals with suspected or confirmed ATP1A3-related disorders (including RDP, AHC, CAPOS) and their family members. It documents both lifetime history and current symptoms, with structured response formats (yes/no, categorical, checklist, text fields). Domains include age and body site of onset, affected regions (including gait, limb and ocular involvement), symptom distribution and temporal course, associated triggers, prior treatments, developmental milestones, episodic events, sensory symptoms, headaches, psychiatric history, behavioral concerns, and potential environmental or physiological triggers associated with symptom onset (**Table 2**). The PHQ also collects information on age at onset, pattern and distribution of initial symptoms, temporal course, and prior diagnostic evaluations, thereby providing contextual clinical data that complements the remote neurological exam (**Table 2**). For individuals with multiple episodes, a supplementary module was used to document the history and phenomenology of up to two additional events.

The Family History Form is a structured tool designed to capture family history for probands with ATP1A3-related disorders (**Table 3**) by collecting standardized information for each family member, including demographics, biological relationship, neurological and psychiatric characteristics, ATP1A3-related phenotypes, and genetic testing results.

Both PHQ and Family History Form were completed by all participants and provided structured background information to support phenotypic characterization.

#### Remote assessment of motor dysfunction via standardized video-based protocol

This fully remote structured neurological examination was adapted from established Dystonia Coalition and Dystonia Study group protocols, with modifications for telemedicine delivery. Our protocol integrated validated motor rating scales ^3,12^, together with cranial nerve screening, speech and swallowing assessment, and functional gait measures, including Timed Up and Go ^13,14^ and 4- and 10-meter walk tasks ^15^ (**Table 4**).

A detailed, standardized manual guided both examiners and participants through all components of the evaluation, specifying camera positioning, lighting, pre-recording instructions, scripted task prompts, and scoring procedures.

The examination assessed motor function, gait, coordination, postural control, and ocular movements using a modified neurological exam that incorporates elements of the Unified Parkinson’s Disease Rating Scale (UPDRS part III), Burke Fahn Marsden Dystonia Scale (BFMDRS), and International Coalition for Ataxia Rating Scale (ICARS) (**Table 4**). Other motor (e.g., finger tapping, toe tapping, hand opening/closing, pronation–supination, leg agility), and speech and voice tasks (see also below) were scored using adapted versions of the same validated rating scales, with defined anchors and criteria for non-performable tasks.

To maximize efficiency and reduce overlap among scales, items were performed in anatomical sequence and grouped by body region, with related assessments conducted consecutively and optimized for remote viewing through standardized camera position.

In addition, the protocol excluded neurological elements that could not be assessed remotely - such as strength, tone, deep tendon reflexes, pupillary responses, and detailed sensory testing. These examination components were omitted for safety or feasibility. In other instances, the protocol provided alternatives or modifications (e.g., adapted heel–knee–shin testing in the seated position, visual observation of eyes closure rather than resistance testing) or included the use of household objects (e.g., a straw for cranial nerve V testing; **Table 4**).

#### Voice and speech analysis

To characterize speech and voice dysfunction, particularly relevant in RDP, in addition to tasks covered in tools like UPDRS, ICARS, BFMDRS previously employed by our group for in person visits ^16,17^, new targeted acoustic and perceptual measures were introduced (**Table 4**). These included the Alternating Motion Rate (AMR), Sequential Motion Rate (SMR) ^18^, and the Speech Intelligibility Test (SIT) ^19–21^. The latter brings together both sentence and word features of scales that have been used extensively to measure the speech intelligibility of persons with motor speech disorders (Assessment of Intelligibility of Dysarthric Speech ^22^, Computerized Assessment of Intelligibility of Dysarthric Speech, CAIDS ^23^ and Phoneme Intelligibility Test ^24^). Standard procedures for the SIT were followed, which involved providing participants with a printed list of 11 sentences ranging in length from 5 to 15 words generated using the SIT software. Speakers were instructed to read aloud the 11 SIT sentences at their comfortable rate and volume. During this data collection session, speakers also read aloud the Rainbow ^25^ and Caterpillar ^26^ Passages at their comfortable rate and volume. When children (<12 y old) were assessed, age-appropriate speech tasks were drawn from the Test of Children’s Speech (TOCS) ^27,28^.

Speech and voice samples were recorded via an H1n portable voice recorder with standardized microphone placement and settings. Recordings were saved as high-quality WAV files.

Self-reported measures included the Voice Handicap Index (VHI-10 ^29^), the Eating Assessment Tool (EAT-10 ^30^) for swallowing-related symptoms, and the Communicative Participation Item Bank (CPIB) ^31^ for assessing the impact of communication difficulties on daily functioning.

#### Neurocognitive and psychological remote testing

Screened participants were scheduled for 1-2 teleneuropsychology appointments (**Table 5**). A comprehensive, age-specific neuropsychological battery (**Table 5**) was administered remotely across one (1.5-2.5 hours), or two sessions (scheduled on a separate date to reduce the effects of fatigue, 1-2 hours), with modifications implemented when necessary for children or individuals requiring additional support. There was one scheduled 5–10-minute break after approximately each hour of testing, with additional breaks available upon request by the participant. Participant responses during the evaluation were documented on digital record forms that were created in both English and Spanish, depending on the participants dominant language. All aspects of a standard neuropsychological evaluation were completed, including clinical interview with participant and caregiver as needed, emotional status assessment, and cognitive testing via telehealth.

The neuropsychological tests that were selected have established reliability and validity in movement disorder populations. Verbal tests that were administered via telehealth did not affect standard administration. For tests with visual components, the examiner used the “Share Screen” feature to present stimulus materials to the participant. The neuropsychological battery covered the gamut of cognitive functions. Intelligence was assessed with the Wechsler Intelligence Scale for Children–4^th^ Edition (WISC-IV) ^32^ or the Wechsler Adult Intelligence Scale-4^th^ Edition (WAIS-IV) ^33^, depending on age. We utilized two common tests of expressive language including the Boston Naming Test (BNT) ^34^ which requires the naming of visually presented objects and the Controlled Oral Word Association Test (COWAT) ^35,36^ which measures the number of words that can be generated in 60 seconds based on a prompt (i.e., letter, semantic category). For auditory/verbal learning and memory, we administered the Word List Learning and Story Memory subtests from the Wide Range Assessment of Memory and Learning- 2^nd^ Edition (WRAML-2) ^37^. Both include assessment of initial learning and retention over 25 minutes. The Benson Figure was used to assess visual/spatial construction and visual memory ^38^.

The battery also included executive function tests including simple attention, working memory, and conceptual reasoning. A digit Span Memory test was derived from the WISC-IV and WAIS-IV. An oral response version of the Trail Making Test ^39,40^ Part A requires visual-motor speed and attention, and Part B adds demand for mental flexibility as patients much switch between two sequences while maintaining visual search and motor control. For cognitive processing speed, the Symbol Digit Modalities Test (SDMT) ^41^, which has shown to be reliably administered orally in a telemedicine environment ^42,43^, was administered in a written (in person) or oral (telehealth) response format. A computerized version of the Wisconsin Card Sorting Test ^44,45^ was administered to assess untimed, conceptual reasoning and cognitive flexibility.

The clinical deficits in this population vary greatly, from those with minimal speech and/or motor deficits to those who are essentially nonverbal and/or unable to write. Tests with no demand for speech with a very low floor were selected for such patients. These tests include the Odd-Item Out subtest from the Reynolds Intellectual Assessment Scales (RIAS2) ^46^, the NEPSY-II Comprehension of Instructions ^47^, and the Test of Visual Perceptual Skills (TVPS) ^48^ Sequential Memory subtest. These are standardized, validated tools that enable us to provide level of performance metrics (e.g., average, below average) for nearly all participants. The tests measure auditory comprehension, working memory, visual episodic memory, and conceptual reasoning. All three tests require only a simple pointing response. Time to completion was captured in all tests. If needed, caregivers stood behind examinees and signaled to the examiner which stimulus response was selected.

Psychiatric scales were also administered. Symptom specific interviews were utilized including the Hamilton Depression Inventory (HDI)^49^ for depression, the Hamilton Anxiety Inventory (HAI) ^8^ for generalized anxiety disorder and the Yale-Brown Obsessive Compulsive Scale (Y-BOCS)^50^ for obsessive-compulsive disorder symptoms. For self-reported symptoms of depression, a mood assessment scale was derived from the Geriatric Depression Scale (GDS) ^51^ for ages ≥18 years, and the Children’s Depression Inventory-2 (CDI-2) ^52^ for ages ≤17 years. To assess for functional ability, the Lawton-Brody Instrumental Activities of Daily Living Scale was administered ^53^. Parents or caregivers completed the Behavior Assessment System for Children–2nd Edition (BASC-2) ^54^ for ages ≤25 years and the Behavior Rating Inventory of Executive Function (BRIEF) ^55^ for ages ≤18 years, via an email link sent to a preferred email address. Adult participants completed the BRIEF-A, also via an email link. Diagnostic-level assessments were conducted using the Quick Structured Clinical Interview for DSM-5 disorders (Quick SCID-5) ^56^, tailored to participant age and cognitive ability.

The neuropsychology team held initial trainings to review standard operating procedures and to determine the order of the test battery to ensure standard administration across our sites.

### Genetic characterization

To confirm *ATP1A3* variant, DNA was extracted from saliva samples at the Massachusetts General Hospital using the prepIT.L2P kit (DNAgenotek) and Sanger sequencing of ATP1A3 exons performed as previously described ^3^.

### Outcomes

The primary outcome of interest was feasibility, defined as the proportion of participants who were able to complete each component of the remote assessment battery at baseline and follow-up visits. Because the purpose of this study was to evaluate implementation rather than clinical outcomes, no diagnostic or inferential analyses were performed, except for verbal and ambulatory status, and dystonia severity.

### Data management

Our study leveraged a robust and secure electronic data infrastructure to ensure high-quality data collection, management, and storage. All clinical data were captured using the REDCap electronic data capture system. The platform supports role-based access controls, audit trails, and automated query management, ensuring compliance with regulatory standards and Good Clinical Practice (GCP) guidelines.

### Statistical analysis

This feasibility-focused study employed descriptive statistical methods. Counts, percentages, means (± SD), medians, and ranges were calculated to summarize demographic characteristics and assessment completion rates. When applicable (due to the observational and descriptive nature of this study), statistical comparisons between controls and participants with ATP1A3 variants were performed using a one-way Welch’s t-test, with significance set at p < 0.05.

### Ethical considerations

All participants (or legal guardians) provided written informed consent in accordance with the Declaration of Helsinki and approved by the Institutional Review Boards at the University of Buffalo (UB; IRB #00006379), which served as the single IRB of record for all participating sites, including the University of Miami (UM), University of California Davis (UCD), Massachusetts General Hospital, and Wake-Forest.

Participants were free to withdraw at any time, and all data management procedures complied with HIPAA and institutional requirements. No adverse events associated with the telemedicine procedures were reported.

## RESULTS

### Screening and Recruitment

Among the subjects who entered the screening pipelines (n = 219; **Figure 1**) social media advertising (Twitter, Facebook, Instagram, and LinkedIn) was the predominant recruitment tool for both screening (47%) and enrollment (44%) (**Figure 2**). Family member referrals increased from screening (7%) to enrollment (22%). Some patients were referred by their neurologists based on clinical suspicion of ATP1A3-related syndromes, including RDP, AHC or CAPOS, or due to the discovery of pathogenic variants in the ATP1A3 gene by third party clinical genetic analysis. Physician referrals comprised 11 and 13% across respectively screening and enrollment. Smaller numbers learned about the study through the UB study website (medicine.buffalo.edu/atp1a3h). Referrals from previous years of our study accounted for similar percentages both at screening and enrollment stages (9% and 7% respectively). Conversely, public database referrals (CT.gov) contributed minimally to overall recruitment (2%). Although digital platforms generated the greatest volume of referrals, personal and clinical sources were proportionally more successful in yielding enrolled subjects.

**Figure 2.**
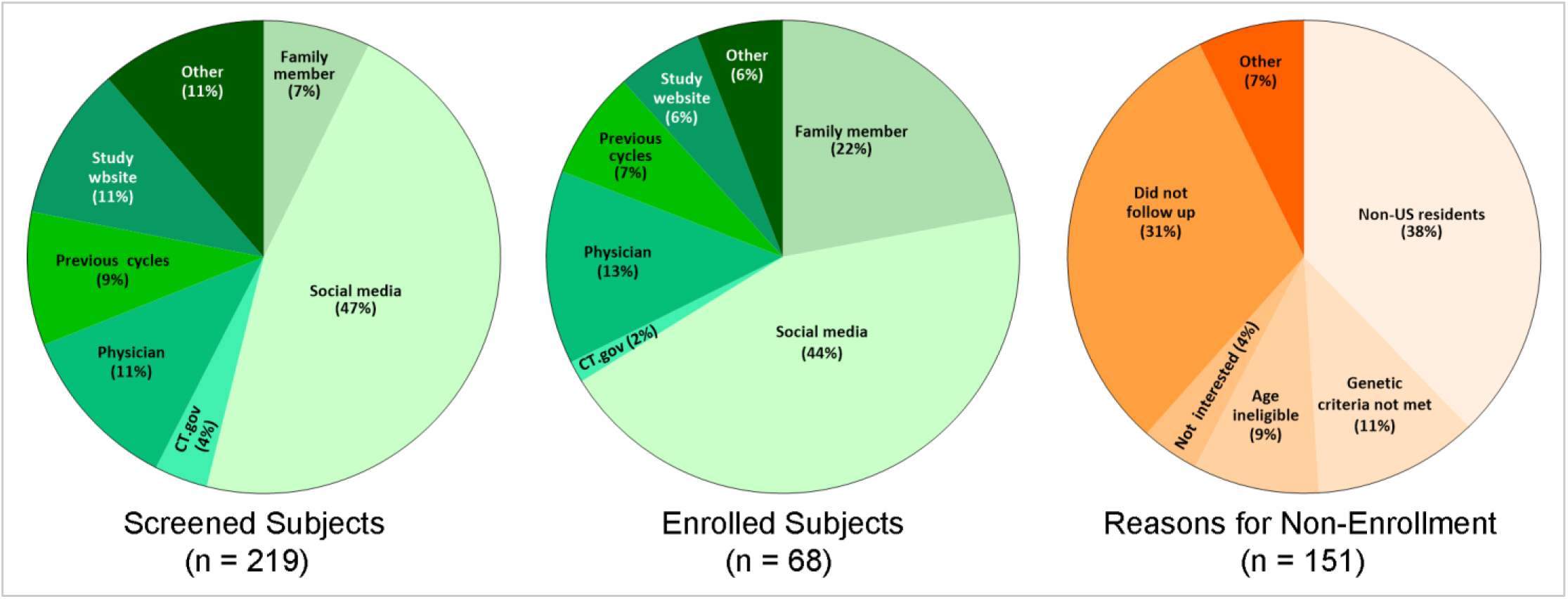
Referral sources for screened and enrolled participants and reasons for non-enrollment. Pie charts illustrating referral sources among screened participants (left) and enrolled participants (center), including social media, family members, physicians, previous study cycles, study website, ClinicalTrials.gov, and other sources. The right panel shows primary reasons for non-enrollment among screened individuals, including non-US residency, lack of follow-up, ineligibility based on age or genetic criteria, lack of interest, and other reasons. Percentages represent the proportion within each category.

Non-enrollment in some cases reflected structural or logistical barriers. More than one-third of individuals were unable to participate because they resided outside the United States, and another 31% did not follow-up after initial contact. Fewer individuals were excluded because they did not meet genetic eligibility criteria (11%), were too young to participate (9%), or declined interest (4%).

The geographic distribution of enrolled participants reflected the breadth and reach of this recruitment approach. Subjects were dispersed widely across the United States, with participants recruited from 23 states spanning the West Coast, Southwest, Midwest, Southeast, and Northeast, with several clusters in regions with active outreach. This broad spatial distribution demonstrates that the fully remote study design enabled participation from individuals who would otherwise be unreachable through traditional site-based enrollment. For the full cohort, the mean time from contact to enrollment was approximately 6 weeks (**Table 6**), with substantial variability. Similar timelines were observed for ATP1A3 cases and control groups. The duration from enrollment to completion averaged approximately 3 months (**Table 6**), reflecting differences in participants availability and scheduling. The time from first to last visit also varied widely, consistent with the longitudinal and flexible nature of the telemedicine protocol.

**Table 6.** Time Intervals from Initial Contact to Study Completion.

|  | Total<br>(n = 56) | ATP1A3<br>(n = 44) | Control<br>(n = 12) |
| --- | --- | --- | --- |
| **Time from Contact to Enrollment (days)** | 43.8 ± 63.1<br>14.5 [0.0, 237.0] | 46.6 ± 66.3<br>14.5 [0.0, 237.0] | 33.5 ± 50.9<br>11.0 [0.0, 160.0] |
| **Time from Enrollment to Completion (days)** | 96.8 ± 94.1<br>65.0 [5.0, 439.0] | 93.0 ± 89.2<br>54.5 [5.0, 350.0] | 110.5 ± 112.7<br>78.0 [7.0, 439.0] |
| **Time from First to Last Visit (days)** | 62.3 ± 82.2<br>31.0 [0.0, 395.0] | 60.5 ± 76.5<br>29.0 [0.0, 322.0] | 68.6 ± 103.5<br>49.0 [0.0, 395.0] |

Data are reported as mean ± SD as well as median [min, max]. Three participants (2 ATP1A3 and 1 control) were excluded due to missing contact dates.

### Feasibility of the telemedicine framework

This telemedicine-adapted neurological examination demonstrated high feasibility at the initial virtual assessment, with a total of 47 participants completing the neurological examination (34 ATP1A3; 13 controls), and only 13 ATP1A3-positive individuals were unable to complete it (**Figure 1**). The speech examination was completed by 40 participants (27 ATP1A3; 13 controls), with 6 ATP1A3 participants not completing this portion. Engagement with the neuropsychological battery showed more variability. Thirty-seven participants completed the full neuropsychological examination (25 ATP1A3; 12 controls). Overall, 22 participants (21 ATP1A3 cases and 1 control) did not complete the neuropsychological battery, reflecting a combination of scheduling challenges and participant burden.

Given the length of the assessments, the neurological/speech exam and neuropsychological testing were performed on different days. For similar reasons, patients were given the option to perform the PHQ and neurological/speech exam on different days.

### Neurological findings

Neurological assessment completion rates were highest among participants with established ATP1A3 phenotypes: 14 of 18 individuals with RDP (78%), 7 of 12 individuals with AHC (58%), 3 of 4 participants with CAPOS (75%), and both “mixed phenotype” cases completed the full examination. In contrast, completion among participants with uncertain ATP1A3 phenotypes (due to unclear age of symptom onset, presence of pathogenic variants without overt clinical manifestations, neurologic features that did not align with established phenotypic categories) was lower, with 4 of 10 individuals (40%) completing the motor exam.

On average, a total of 1.8 (± 0.6; SD) visits were required to complete the entire neurological/speech battery, with statistically significantly more sessions (36%; p = 0.006) necessary for ATP1A3 cases, relative to controls (**Table 8**). Assessment time for the overall neurological session (including speech testing) was (mean ± SD) 55.2 ± 18.0 minutes (**Table 7**), with statistically longer duration (by 30%; p < 0.0001) in ATP1A3 cases relative to controls (**Table 7**), indicating greater clinical complexity among affected individuals.

**Table 7.** Duration of Neurological and Speech Evaluations.

|  | Total<br>(n = 47) | ATP1A3<br>(n = 34) | Control<br>(n = 13) |
| --- | --- | --- | --- |
| **Total Time per Session (min)** | 55.2 ± 18.0<br>54.1 [8.4, 93.6] | 58.9 ± 19.6<br>57.4 [8.4, 93.6] | 45.5 ± 7.0*<br>45.6 [37.1, 59.7] |
| **Neurological Visit (min)** | 46.6 ± 14.4<br>45.0 [8.4, 78.4] | 50.2 ± 15.0<br>50.0 [8.4, 78.4] | 37.0 ± 6.3**<br>37.0 [29.9, 52.4] |
| **Speech Assessment (min)** | 8.6 ± 5.7<br>8.7 [0.0, 27.8] | 8.7 ± 6.6<br>8.9 [0.0, 27.8] | 8.5 ± 1.5<br>7.8 [6.4, 11.4] |

Data are shown as mean ± SD and median [min, max]. Table includes only participants with non-missing data.
\* p = 0.0006 by one-tailed Welch's t-test between controls and ATP1A3 ; \*\* p < 0.0001 by one-tailed Welch's t-test between controls and ATP1A3

**Table 8.** Frequency of Neurological and Neurocognitive Visits.

|  | Total | ATP1A3 | Control |
| --- | --- | --- | --- |
| **Number of Neurological Visits** | 1.8 ± 0.6<br>2.0 [1.0, 3.0]<br>(n = 47) | 1.9 ± 0.6<br>2.0 [1.0, 3.0]<br>(n = 34) | 1.4 ± 0.5*<br>1.0 [1.0, 2.0]<br>(n = 13) |
| **Number of Neurocognitive/Psychological Visits** | 1.0 ± 1.0<br>1.0 [1.0, 3.0]<br>(n = 34)‡ | 1.0 ± 1.1<br>0.0 [1.0, 3.0]<br>(n = 22)‡ | 1.1 ± 0.5<br>1.0 [1.0, 2.0]<br>(n = 12) |

Data are reported as mean ± SD and median [min, max]. Neurological visits also include speech assessments. ‡ To date, 37 participants have completed the neuropsychological evaluation, with data from 3 participants still under analysis. \* p = 0.0029 by one-tailed Welch's t-test between controls and ATP1A3.

Among participants with available gait data, most of ATP1A3 cases retained independent ambulation, with 26 of 32 individuals able to walk (**Table 9**). Preserved gait was most common in participants with AHC (89%) and CAPOS (100%), whereas greater variability was observed among individuals with RDP (81%) and undefined phenotypes (50%). All controls exhibited independent ambulation.

**Table 9.**
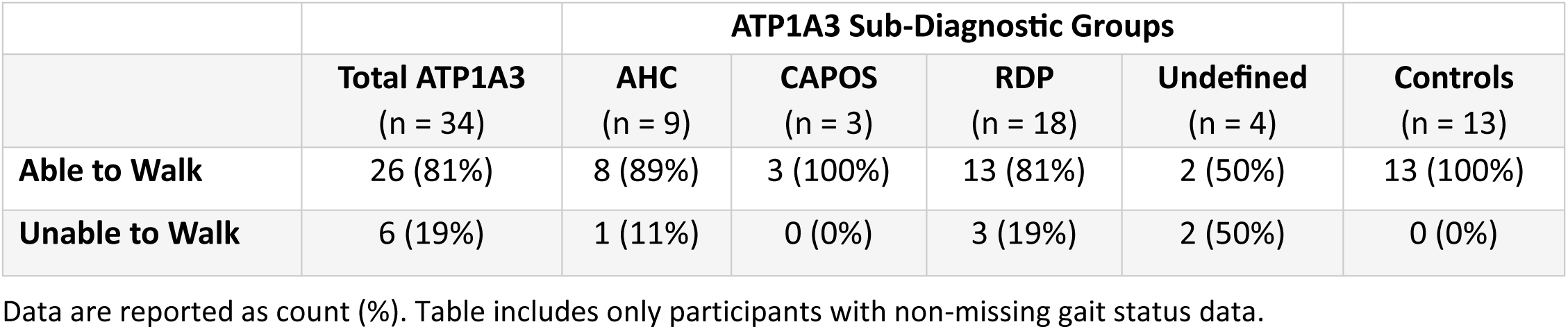
Proportion of Participants Affected by Gait Impairment.

Dystonia severity (extrapolated by BFMDRS scores) differed markedly across diagnostic groups (**Table 10**). Participants with RDP demonstrated the highest mean and median severity scores (55.0 ± 50.0), consistent with prominent and persistent motor involvement in this cohort. In contrast, individuals with AHC and CAPOS exhibited substantially lower dystonia severity (41.0 ± 44.7 and 8.7 ± 2.8 respectively), reflecting milder or more episodic motor manifestations. Overall, all subgroups showed wide variability in severity scores, consistent with the well-known heterogeneous clinical presentation and variable disease burden characterizing the ATP1A3 population ^3,12^. Among controls, one outlier reflected secondary dystonia following neck surgery in a family member of an ATP1A3 case (**Table 10**).

**Table 10.** Dystonia Severity Scores.

|  | ATP1A3 Sub-Diagnostic Groups |  |  |  |  |
| --- | --- | --- | --- | --- | --- |
| **Total ATP1A3**<br>(n = 31) | **AHC**<br>(n = 8) | **CAPOS**<br>(n = 3) | **RDP**<br>(n = 16) | **Undefined**<br>(n = 4) | **Controls**<br>(n = 13) |
| 46.9 ± 47.3 | 41.0 ± 44.7 | 8.7 ± 2.8 | 55.0 ± 50.0 | 53.3 ± 56.3 | 0.1 ± 0.3 |
| 26.5 | 23.5 | 8.5 | 32.0 | 53.0 | 0.0 |
| [3.0, 204.0] | [8.5, 138.5] | [6.0, 11.5] | [7.0, 204.0] | [3.0, 104.0] | [0.0, 1.0<sup>‡</sup>] |

Data are reported as mean ± SD as well as median [min, max]. Table includes only participants with non-missing dystonia severity data.
<sup>‡</sup> Mother of an ATP1A3 case; developed dystonia following neck surgery.

Together, these findings demonstrate that a structured, motor-focused neurological examination and associated gait tasks can be conducted remotely in approximately 78% of enrolled participants (**Figure 1**), including those with complex movement disorders and across diverse phenotypes. Also, these subtype-specific patterns of gait impairment and dystonia severity are consistent with our prior observations from in-person clinical evaluations ^1,3,11,12^, supporting the validity of remote assessments.

### Verbal abilities by diagnostic subgroups

A total of 40 participants (all 13 controls and 27 ATP1A3; 68%) completed the speech exam, while 6 individuals with ATP1A3 variants did not (**Figure 1**). Also in this instance, the higher compliance was found among individuals with RDP, AHC and CAPOS (93%, 71% and 100% respectively) ^16^, whereas of the 10 subjects with undefined phenotype, only 2 completed the voice/speech exam.

The mean duration of speech evaluations was 8.6 (± 5.7) minutes for the full cohort, with similar times observed for ATP1A3 participants (8.7 ± 6.6 minutes) and controls (8.5 ± 1.5 minutes) (**Table 7**). Unlike neurological examinations, the similar speech assessment duration suggests comparable feasibility and efficiency of remote speech evaluation across diagnostic categories (**Table 7**).

Verbal ability was largely preserved across our cohort (**Table 11**). Among participants with available data, the majority were able to speak, including most individuals with established ATP1A3 phenotypes (81%) and controls (100%). High rates of intact verbal abilities were observed among individuals with AHC (88%) and CAPOS (100%). Participants with RDP also largely retained verbal ability (81%), although a small subset (19%) exhibited reduced speech. In contrast, reduced verbal ability was more frequent among individuals with undefined phenotypes.

**Table 11.** Verbal Status Across Diagnostic Groups.

|  |  | ATP1A3 Sub-Diagnostic Groups |  |  |  |  |
| --- | --- | --- | --- | --- | --- | --- |
|  | **Total ATP1A3**<br>(n = 33) | **AHC**<br>(n = 8) | **CAPOS**<br>(n = 3) | **RDP**<br>(n = 18) | **Undefined**<br>(n = 4) | **Controls**<br>(n = 13) |
| **Able to Speak** | 25 (81%) | 7 (88%) | 3 (100%) | 13 (81%) | 2 (50%) | 13 (100%) |
| **Unable to Speak** | 6 (19%) | 1 (13%) | 0 (0%) | 3 (19%) | 2 (50%) | 0 (0%) |

Data are reported as count (%). Table includes only participants with non-missing verbal status data.

Overall, patterns of speech impairments across diagnostic subgroups were consistent with prior in-person evaluations^16^, further supporting the validity of remote speech assessments and highlighting subtype-specific differences in communicative function.

### Neuropsychological assessment

Neuropsychological assessment was completed over 1-2 sessions for controls and 1-3 sessions for individuals with ATP1A3 variants (**Table 8**). A total of 37 participants (25 ATP1A3 variant carriers and 12 controls) were evaluated. Some participants were unable to tolerate lengthy cognitive testing. Accordingly, the goal of comprehensive data collection was balanced with the need to minimize participant burden and maintain engagement. Priority was therefore given to ensuring completion of the core cognitive battery whenever possible.

Analyses are currently ongoing; however, initial findings indicate that the effect size comparing ATP1A3 cases and controls are greatest in the memory domain, including word list learning (d = 2.8) and narrative text recall (d = 2.8; p < 0.001). In contrast, the smallest group differences were observed for estimated verbal IQ (d = 0.2; p = 0.60), suggesting relatively preserved performance in this domain.

These early results highlight and confirm a marked variability in cognitive profiles ^3,12^ among ATP1A3 variant carriers and underscore the particular vulnerability of memory-related function in this population.

## DISCUSSION

The current study demonstrates the feasibility of implementing a comprehensive telemedicine-based assessment protocol for the phenotyping of RDP.

Building on our prior work, which identified substantial phenotypic variability both longitudinally and across individuals ^3,12^, we extended our patient outreach by integrating remote neurological, neurocognitive and voice assessment into a unified telehealth platform. This framework not only enhances access and at-home characterization of individuals with rare ATP1A3 disorders but can also be adapted to the evaluation of a broader range of neurological diseases that share overlapping symptoms (e.g., other dystonias, hereditary and sporadic ataxia, mitochondrial-related movement disorders, myasthenia gravis, etc). In agreement with this, telemedicine has already been shown to be feasible and reliable in movement disorders such as Parkinson’s disease (PD), dystonia ^57^, and Huntington’s disease ^8^. The same integrated approach could benefit patients with neurogenetic and neurodevelopmental conditions (e.g., FXTAS, Rett syndrome).

The adoption of telemedicine represents a key methodological advance for the study of RDP. By conducting interviews and standardized assessments in patients’ familiar environments, we can capture real-world data on symptom severity and progression. Remote blinded examinations and structured video-based ratings, modeled after protocols from the Dystonia Coalition, ensure diagnostic consistency while minimizing observer bias. Importantly this approach enables longitudinal monitoring and data harmonization across sites.

Prior studies from our group ^3,12,16^ have shown that classic diagnostic criteria (e.g., rapid onset and rostral-caudal distribution) do not encompass the full variability of ATP1A3-related presentations. Compared to that body of work, the remote framework facilitated the recruitment and longitudinal assessment of participants across a broad clinical spectrum, including those with atypical or milder phenotypes.

Our expanded battery incorporates objective, scalable and telemedicine-compatible tools spanning neurological, neurocognitive and behavioral (voice/speech) domains. The use of teleneuropsychology is particularly important for individuals with ATP1A3-related disorders who often experience episodic or progressive motor dysfunction, gait instability, fatigue and bulbar symptoms that can make travel to specialized centers physically demanding and unsafe. Remote assessment minimizes the need for long commutes, decreases fall risk, reduces participant fatigue, and allows for increased involvement of caregivers for optimal care. The introduction of neuropsychiatric and self-report instruments, including the VHI-10 and EAT-10 questionnaires, allows detection of psychiatric, cognitive and episodic bulbar manifestations that may otherwise go unreported. The inclusion of structured voice recordings and blinded acoustic ratings further refine our previous characterization of dysarthria and dysphonia ^3,16^, features that are now recognized as common and clinically relevant in ATP1A3-related disease. These measures not only improve diagnostic precision but also hold potential as quantifiable biomarkers for disease monitoring as well as therapeutic response.

In this initial analysis, we focused on a limited set of core functional domains—dystonia severity, verbal capacity, and gait function—to provide a preliminary characterization of motor and communicative outcomes within this large, deeply phenotyped ATP1A3 cohort. These domains were selected because they represent clinically meaningful indicators of disease burden, are reliably assessable in a remote setting, and are central to patient quality of life. Despite this narrow analytic scope, clear subtype-specific patterns emerged, with greater motor impairment in individuals with RDP, preserved speech in most participants, and variable gait dysfunction across phenotypes, consistent with prior in-person observations.

Importantly, these findings demonstrate the feasibility of extracting robust, clinically relevant measures from a large, geographically distributed telemedicine dataset. The present analyses represent an initial step toward comprehensive longitudinal modeling of disease trajectories, treatment response, and genotype–phenotype relationships. Ongoing and future work will extend this framework to incorporate additional neurological, cognitive, behavioral, and environmental variables captured in this cohort, enabling more refined characterization of disease progression and heterogeneity in ATP1A3-related disorders.

Further iterations of our protocol may include automated video and speech analysis algorithms to enhance objectivity and scalability. In addition, recent advances in AI and computer vision have paved the way toward new possibilities for remote quantification of oculomotor functions. AI-based eye-tracking tools and smartphone compatible video analytics can now extract fine parameters of gaze stability and saccade velocity from standard video recordings. Lastly, integrating remotely acquired but readily shared ECG data will allow for a more comprehensive analysis of the ATP1A3 pathology ^58^.

### Limitations

Although our protocol provides a comprehensive and valid remote assessment tool, several standard neurological examination elements cannot be performed or need to be modified due to telemedicine constraints. With the limited ability to distinguish very subtle phenomena via video, some features may be more difficult to reliably quantify, such as mild rigidity, paratonia, or spasticity; and subtle oculomotor abnormalities. In addition, this protocol relies heavily on patient/caregiver home environment, and task performance may be affected by a number of variables like: (i) space for walking tasks (e.g., narrow halls, cluttered rooms), (ii) flooring type (carpet vs. hardwood), (iii) lighting and camera angle that might obscure tremor, postural changes, or foot placement, and (iv) internet bandwidth. We recognize other limitations that may also occur with telemedicine, including lack of technological equipment, discomfort with remote testing, or issues of confidentiality.

## Conclusions

Our findings support telemedicine as a powerful, patient-centered approach for RDP phenotyping. By merging neurological, neurocognitive and voice assessments into a unified remote framework, we expand both the reach and resolution of clinical characterization. This approach not only enhances care access and inclusivity but also lays the foundation for long-term longitudinal studies, digital biomarkers discovery, and the development of targeted therapies for ATP1A3-related movement disorders.

## Acknowledgments

We gratefully acknowledge Dr. Beverly Snively for developing the Wake-Forest patient database used during the initial phase of this study. We also extend our sincere appreciation to the participating individuals and their families. Their commitment and generosity made this work possible.

## Data Availability

A partial deidentified dataset is available upon request.

## Authors’ Contribution

IUH, DS, VLW, and AB developed the neurological assessment protocol, conducted neurological evaluations, and participated in diagnostic consensus meetings. RHB and MLS developed telemedicine neuropsychological assessment battery and conducted participant testing. KT contributed to the design of the voice and speech assessment. LJO oversaw genetic testing. RF coordinated and scheduled neurological assessments at UB, managed participant communication, and performed data entry and extraction. EN drafted the manuscript, prepared figures and tables, and performed statistical analyses. KJS provided critical input on manuscript development and edited the manuscript. AB conceived and led the study. All authors reviewed and approved the final version of the manuscript.

## REFERENCES

1 Brashear, A., Dobyns, W. B., de Carvalho Aguiar, P. et al. The phenotypic spectrum of rapid-onset dystonia-parkinsonism (RDP) and mutations in the *ATP1A3* gene. Brain 130, 828–835 (2007). doi:10.1093/brain/awl340

2 Dobyns, W. B., Ozelius, L. J., Kramer, P. L. et al. Rapid-onset dystonia-parkinsonism. Neurology 43, 2596–2602 (1993). doi:10.1212/wnl.43.12.2596

3 Haq, I. U., Snively, B. M., Sweadner, K. J. et al. Revising rapid-onset dystonia-parkinsonism: Broadening indications for ATP1A3 testing. Mov Disord 34, 1528–1536 (2019). doi:10.1002/mds.27801

4 Heinzen, E. L., Arzimanoglou, A., Brashear, A. et al. Distinct neurological disorders with ATP1A3 mutations. Lancet Neurol 13, 503–514 (2014). doi:10.1016/S1474-4422(14)70011-0

5 Rosewich, H., Ohlenbusch, A., Huppke, P. et al. The expanding clinical and genetic spectrum of ATP1A3-related disorders. Neurology 82, 945–955 (2014). doi:10.1212/WNL.0000000000000212

6 Vezyroglou, A., Akilapa, R., Barwick, K. et al. The phenotypic continuum of ATP1A3-related disorders. Neurology 99, e1511–e1526 (2022). doi:10.1212/WNL.0000000000200927

7 Larner, A. J. Teleneurology: An overview of current status. Pract Neurol 11, 283–288 (2011). doi:10.1136/practneurol-2011-000090

8 Hatcher-Martin, J. M., Adams, J. L., Anderson, E. R. et al. Telemedicine in neurology: Telemedicine Work Group of the American Academy of Neurology update. Neurology 94, 30–38 (2020). doi:10.1212/WNL.0000000000008708

9 Dorsey, E. R., Glidden, A. M., Holloway, M. R., Birbeck, G. L. & Schwamm, L. H. Teleneurology and mobile technologies: The future of neurological care. Nat Rev Neurol 14, 285–297 (2018). doi:10.1038/nrneurol.2018.31

10 Lewinski, A. A., Walsh, C., Rushton, S. et al. Telehealth for the longitudinal management of chronic conditions: Systematic review. J Med Internet Res 24, e37100 (2022). doi:10.2196/37100

11 Brashear, A., Sweadner, K. J., Haq, I., Napoli, E. & Ozelius, L. in GeneReviews(R), eds M. P. Adam et al. (1993 [Updated 2024, Dec 5]).

12 Haq, I. U., Napoli, E., Snively, B. M. et al. Neurological and psychiatric characterization of rapid-onset dystonia-parkinsonism over time. Parkinsonism Relat Disord 131, 107254 (2025). doi:10.1016/j.parkreldis.2024.107254

13 Podsiadlo, D. & Richardson, S. The timed “Up & Go“: A test of basic functional mobility for frail elderly persons. J Am Geriatr Soc 39, 142–148 (1991). doi:10.1111/j.1532-5415.1991.tb01616.x

14 Beauchet, O., Fantino, B., Allali, G. et al. Timed Up and Go test and risk of falls in older adults: A systematic review. J Nutr Health Aging 15, 933–938 (2011). doi:10.1007/s12603-011-0062-0

15 Bohannon, R. W. & Wang, Y. C. Four-Meter Gait Speed: Normative values and reliability determined for adults participating in the NIH Toolbox study. Arch Phys Med Rehabil 100, 509–513 (2019). doi:10.1016/j.apmr.2018.06.031

16 Moya-Mendez, M. E., Madden, L. L., Ruckart, K. W. et al. in J Clin Neurosci Vol. 81 133–138 (2020).

17 Duffy, J. R. Motor speech disorders: Substrates, differential diagnosis, and management 4th edn, Elsevier, (2019).

18 Pierce, J. E., Cotton, S. & Perry, A. Alternating and sequential motion rates in older adults. Int J Lang Commun Disord 48, 257–264 (2013). doi:10.1111/1460-6984.12001

19 Stipancic, K. L. & Tjaden, K. Minimally detectable change of speech intelligibility in speakers with Multiple Sclerosis and Parkinson’s Disease. J Speech Lang Hear Res 65, 1858–1866 (2022). doi:10.1044/2022_JSLHR-21-00648

20 Stipancic, K. L., Wilding, G. & Tjaden, K. Lexical characteristics of the Speech Intelligibility Test: Effects on transcription intelligibility for speakers with Multiple Sclerosis and Parkinson’s Disease. J Speech Lang Hear Res 66, 3115–3131 (2023). doi:10.1044/2023_JSLHR-22-00279

21 Yorkston, K. M., Beukelman, D. & Hakel, M. Speech Intelligibility Test (SIT) for Windows [computer software]. Madonna Rehabilitation Hospital. (2007).

22 Yorkston, K. M. & Beukelman, D. R. Communication efficiency of dysarthric speakers as measured by sentence intelligibility and speaking rate. J Speech Hear Disord 46, 296–301 (1981). doi:10.1044/jshd.4603.296

23 Yorkston, K. M., Beukelman, D. R. & Traynor, C. Computerized assessment of intelligibility of dysarthric speech. Tigard, Ore. : C.C. Publications, (1984).

24 Van Nuffelen, G., Middag, C., De Bodt, M. & Martens, J. P. Speech technology-based assessment of phoneme intelligibility in dysarthria. Int J Lang Commun Disord 44, 716–730 (2009). doi:10.1080/13682820802342062

25 Fairbanks, G. in Voice and Articulation Drillbook, Harper Bros, (1960).

26 Patel, R., Connaghan, K., Franco, D. et al. “The caterpillar“: A novel reading passage for assessment of motor speech disorders. Am J Speech Lang Pathol 22, 1–9 (2013). doi:10.1044/1058-0360(2012/11-0134)

27 Hodge, M., J. Daniels & Gotzke., C. L. TOCS+ intelligibility measures. University of Alberta, (2007).

28 Hodge, M. M. & Gotzke, C. L. Construct-related validity of the TOCS measures: Comparison of intelligibility and speaking rate scores in children with and without speech disorders. J Commun Disord 51, 51–63 (2014). doi:10.1016/j.jcomdis.2014.06.007

29 Rosen, C. A., Lee, A. S., Osborne, J., Zullo, T. & Murry, T. Development and validation of the Voice Handicap Index-10. Laryngoscope 114, 1549–1556 (2004). doi:10.1097/00005537-200409000-00009

30 Belafsky, P. C., Mouadeb, D. A., Rees, C. J. et al. Validity and reliability of the Eating Assessment Tool (EAT-10). Ann Otol Rhinol Laryngol 117, 919–924 (2008). doi:10.1177/000348940811701210

31 Baylor, C., Yorkston, K., Eadie, T. et al. The Communicative Participation Item Bank (CPIB): Item bank calibration and development of a disorder-generic short form. J Speech Lang Hear Res 56, 1190–1208 (2013). doi:10.1044/1092-4388(2012/12-0140)

32 Wechsler, D. Weschler Intelligence Scale for Children - Fourth Edition, Administration and Scoring Manual. The Psychological Corporation, (2003).

33 Wechsler, D. Wechsler Adult Intelligence Scale, Fourth Edition. Pearson, (2008).

34 Kaplan, E. F., Goodglass, H. & Weintraub, S. The Boston Naming Test. Lea & Febiger, (1983).

35 Ruff, R. M., Light, R. H., Parker, S. B. & Levin, H. S. Benton Controlled Oral Word Association Test: Reliability and updated norms. Archives of Clinical Neuropsychology 11, 329–338 (1996).

36 Borkowski, J. G., Benton, A. L. & Spreen, O. Word fluency and brain damage. Neuropsychologia 5, 135–140 (1967).

37 Sheslow, D. a. A., W. Wide range assessment of memory and learning. Second edition. Wide Range Inc, (2003).

38 Dodge, H. H., Goldstein, F. C., Wakim, N. I. et al. Differentiating among stages of cognitive impairment in aging: Version 3 of the Uniform Data Set (UDS) neuropsychological test battery and MoCA index scores. Alzheimers Dement (N Y) 6, e12103 (2020). doi:10.1002/trc2.12103

39 Heaton, R. K., Grant, I. & Matthews, C. G. Comprehensive norms for an expanded Halstead-Reitan Battery: Demographic corrections, research findings, and clinical applications. Psychological Assessment Resources, (1991).

40 Reitan, R. M. Validity of the Trail Making Test as an indicator of organic brain damage. Perceptual and Motor Skills 8, 271–276 (1958).

41 Smith, A. Symbol digit modalities test: Manual. Western Psychological Services, (1982).

42 Eilam-Stock, T., Shaw, M. T., Sherman, K., Krupp, L. B. & Charvet, L. E. Remote administration of the symbol digit modalities test to individuals with multiple sclerosis is reliable: A short report. Mult Scler J Exp Transl Clin 7, 2055217321994853 (2021). doi:10.1177/2055217321994853

43 Levy, S., Dvorak, E. M., Graney, R., Staker, E. & Sumowski, J. F. In-person and remote administrations of the symbol digit modalities test are interchangeable among persons with multiple sclerosis. Mult Scler Relat Disord 71, 104553 (2023). doi:10.1016/j.msard.2023.104553

44 Heaton, R. K. A Manual for the Wisconsin Card Sorting Test. Psychological Assessment Resources, Inc., (1981).

45 Arnett, P. A., Rao, S. M., Bernardin, L. et al. Relationship between frontal lobe lesions and Wisconsin Card Sorting Test performance in patients with multiple sclerosis. Neurology 44, 420–425 (1994).

46 Reynolds, C. R., & Kamphaus, R. W. Reynolds Intellectual Assessment Scales, Second Edition (RIAS-2) professional manual. Psychological Assessment Resources, (2015).

47 Korkman, M., Kirk, U. & Kemp, S. L. NEPSY–II: Clinical and Interpretive Manual. Harcourt Assessment, (2007).

48 Martin, N. A. Test of Visual Perceptual Skills, Fourth Edition (TVPS-4) manual. Academic Therapy Publications, (2017).

49 Kobak, K. A. & Reynolds, W. M. in The use of psychological testing for treatment planning and outcomes assessment: Instruments for adults eds M. E. Maruish 327–362 Lawrence Erlbaum Associates Publishers, (2004).

50 Goodman, W. K., Price, L. H., Rasmussen, S. A. et al. The Yale-Brown Obsessive Compulsive Scale. I. Development, use, and reliability. Arch Gen Psychiatry 46, 1006–1011 (1989). doi:10.1001/archpsyc.1989.01810110048007

51 Ferraro, F. R. & Chelminski, I. Preliminary normative data on the geriatric depression scale-short form (GDS-SF) in a young adult sample. J Clin Psychol 52, 443–447 (1996). doi:10.1002/(SICI)1097-4679(199607)52:4<443::AID-JCLP9>3.0.CO;2-Q

52 Kovacs, M. in The Encyclopedia of Clinical Psychology, eds R.L. Cautin and S.O. Lilienfeld Multi-Health Systems, Inc, (2011).

53 Lawton, M. P. & Brody, E. M. Instrumental Activities of Daily Living Scale. APA PsycTests. (1969).

54 Reynolds, C. R. & Kamphaus, R. W. Behavior assessment system for children - Second edition (BASC-2). American Guidance Service, (2004).

55 Gioia, G. A., Isquith, P. K., Guy, S. C. & Kenworthy, L. Behavior rating of executive function. Psychological Assessment Resources, (2000).

56 First, M. B. & Williams, J. B. W. Quick Structured Clinical Interview for DSM-5 Disorders (QuickSCID-5). 1–61 American Psychiatric Association Publishing, (2020).

57 Bragg, D. C., Sharma, N. & Ozelius, L. J. X-Linked Dystonia-Parkinsonism: Recent advances. Curr Opin Neurol 32, 604–609 (2019). doi:10.1097/WCO.0000000000000708

58 Moya-Mendez, M. E., Bidzimou, M. T., Muralidharan, P. et al. ATP1A3 variants, variably penetrant short QT intervals, and lethal ventricular arrhythmias. JAMA Pediatr 179, 529–539 (2025). doi:10.1001/jamapediatrics.2024.6832

